# MedScope: A Lightweight Benchmark of Open-Source Large Language Models for Medical Question Answering

**DOI:** 10.64898/2026.03.31.26349827

**Authors:** Rui Bian, Weihao Cheng

## Abstract

The rapid development of large language models (LLMs) has stimulated growing interest in their use for medical question answering and clinical decision support. However, compared with frontier proprietary systems, the empirical understanding of lightweight open-source LLMs in medical settings remains limited, particularly under resource-constrained experimental conditions. To address this gap, we introduce MedScope, a lightweight benchmarking framework for systematically evaluating open-source LLMs on medical multiple-choice question answering.

Using 1,000 sampled questions from MedMCQA, we benchmark six lightweight open-source models spanning three representative model families: LLaMA, Qwen, and Gemma. Beyond standard predictive metrics such as accuracy and macro-F1, our framework additionally considers inference time, prediction consistency, subject-wise variability, and model-specific error patterns. We further develop a set of multi-perspective visual analyses, including clustered heatmaps, agreement matrices, Pareto-style trade-off plots, radar charts, and multi-panel summary figures, in order to characterize model behavior in a more interpretable and comprehensive manner.

Our results reveal substantial heterogeneity across models in predictive performance, efficiency, and subject-level robustness. While larger lightweight models generally achieve better overall results, the gain is neither uniform across subject categories nor always aligned with efficiency. These findings suggest that lightweight open-source LLMs remain valuable as transparent and reproducible medical AI baselines, but their current capabilities are still insufficient for unsupervised deployment in high-risk healthcare scenarios. MedScope provides an accessible benchmark for evaluating lightweight medical LLMs and emphasizes the need for multi-dimensional assessment beyond accuracy alone.The relevant code is now open-sourced at: https://github.com/VhoCheng/MedScope.

## 1 Introduction

Large language models (LLMs) have recently demonstrated strong capabilities in knowledge-intensive language tasks, leading to increasing interest in their use for medical question answering, clinical decision support, and healthcare education [8,10,11]. In particular, medical question answering has emerged as an important testbed for evaluating whether LLMs can retrieve, reason over, and communicate domain-specific knowledge in a clinically meaningful manner [9, 11]. Nevertheless, despite rapid progress in frontier proprietary systems, the behavior of lightweight open-source LLMs in medical scenarios remains comparatively underexplored.

This gap is important for both scientific and practical reasons. Compared with closed commercial systems, lightweight open-source LLMs offer advantages in accessibility, transparency, reproducibility, and local deployment, which are highly relevant for academic benchmarking and resource-constrained healthcare settings [5, 8, 16]. Such models make it possible to conduct controlled and reproducible experiments without relying on remote proprietary APIs, while also reducing barriers to deployment in settings where computational resources, privacy constraints, or budget limitations make large-scale commercial systems impractical [8]. However, these benefits do not automatically imply sufficient medical competence, and systematic evaluation is therefore essential.

Existing medical QA benchmarks have provided an important foundation for this line of research. Among them, MedMCQA is one of the most widely used large-scale multi-subject medical multiple-choice datasets, containing questions spanning diverse medical disciplines and requiring broad biomedical knowledge [9]. Prior work on medical LLMs has shown that scale can substantially improve performance, as illustrated by Med-PaLM and subsequent expert-level medical QA studies [10, 11]. At the same time, the literature has repeatedly emphasized that medical LLM assessment should not rely on a single headline score alone, because issues such as robustness, explainability, reliability, and deployment risk remain central in healthcare applications [2, 8].

In this context, lightweight model benchmarking remains especially necessary. First, current studies disproportionately emphasize large or proprietary models, while the practical performance ceiling of smaller open-source models is still unclear. Second, many evaluations focus primarily on overall correctness, but provide limited analysis of efficiency, inter-model agreement, and subject-wise variability. Third, in high-stakes domains such as medicine, a model that appears acceptable on aggregate accuracy may still behave unevenly across specialties or exhibit unstable prediction patterns, which weakens confidence in downstream use [8, 11]. Therefore, a more comprehensive and interpretable evaluation framework is needed for lightweight medical LLMs.

To address this gap, we present **MedScope**, a lightweight benchmark for evaluating open-source LLMs on medical multiple-choice question answering. Using 1,000 sampled questions from MedMCQA, we systematically compare six lightweight models from three representative model families, namely LLaMA, Qwen, and Gemma, under a unified prompting and evaluation protocol. Beyond conventional metrics such as accuracy and macro-F1, we further examine inference efficiency, invalid response rate, inter-model agreement, subject-wise performance differences, and several visual trade-off patterns. In this way, our study aims to move beyond single-metric comparison and provide a more multi-dimensional understanding of how lightweight open-source LLMs behave in medical QA.

The main contributions of this work are threefold. First, we establish a lightweight and reproducible evaluation setting for medical multiple-choice question answering using open-source LLMs. Second, we conduct a systematic comparison across multiple lightweight model families and parameter scales. Third, we provide an interpretable multi-view analysis of performance, efficiency, and consistency, offering practical evidence for the strengths and limitations of lightweight medical LLMs in resource-constrained settings.

## 2 Related Work

### 2.1 Medical Large Language Models in Healthcare

The rapid progress of large language models (LLMs) has stimulated broad interest in their application to healthcare, including medical question answering, knowledge retrieval, patient education, diagnostic assistance, and clinical dialogue. Early landmark work such as Med-PaLM demonstrated that general-purpose LLMs could be adapted to the medical domain and evaluated on diverse medical QA benchmarks, while later studies further pushed performance toward expert-level medical question answering [10, 11]. More recently, conversational systems such as AMIE have extended this line of research from static question answering to diagnostic dialogue, highlighting the growing ambition of medical LLMs in real-world healthcare interaction [14].

At the same time, the literature has made it increasingly clear that strong performance on benchmark-style medical tasks does not automatically imply readiness for clinical use. Reviews and perspective papers have consistently emphasized that medical LLMs should be evaluated not only for factual correctness, but also for reliability, safety, robustness, fairness, privacy, and explainability [1, 2, 8]. This concern is particularly important in healthcare, where seemingly plausible but incorrect responses can produce misleading advice, obscure responsibility boundaries, and amplify downstream risk.

### 2.2 Medical Question Answering Benchmarks and Evaluation Paradigms

Medical question answering has become one of the most widely used testbeds for assessing domain-specific capabilities of LLMs. A central milestone in this area is MultiMedQA, which aggregates several existing medical QA benchmarks spanning professional medicine, consumer health, and biomedical research, thereby encouraging broader and more realistic evaluation [10,11]. Within the multiple-choice setting, MedMCQA has emerged as one of the most influential benchmarks, providing a large-scale, multi-subject dataset of medical entrance-exam-style questions covering diverse disciplines and topics [9]. Because of its scale and disciplinary coverage, MedMCQA is particularly suitable for comparing model behavior across specialties rather than relying only on a single aggregate score.

Recent work has also begun to highlight limitations in conventional evaluation practices. Several studies note that medical LLM assessment often overemphasizes final answer correctness while under-analyzing inter-model consistency, efficiency, distributional differences across subject areas, and error structure [3, 7, 8]. In parallel, benchmark-oriented studies in biomedical NLP have shown that model performance can vary substantially across task types and application settings, reinforcing the importance of more fine-grained and multi-dimensional assessment strategies [4]. These observations motivate benchmarking frameworks that move beyond headline accuracy and incorporate richer behavioral analysis.

### 2.3 Lightweight Open-Source Models and Reproducible Evaluation

Although proprietary frontier models have driven many of the headline advances in medical LLM research, lightweight open-source models are increasingly important for reproducible experimentation, local deployment, and low-resource research settings. Open technical reports such as Llama 3, Qwen2.5, and Gemma/Gemma 3 document the rapid improvement of open-weight models across reasoning, instruction following, multilinguality, and efficiency-oriented design [5, 12, 13, 16]. These model families provide an increasingly strong foundation for studying medical QA without relying on opaque remote APIs.

However, despite their practical relevance, lightweight open-source LLMs remain under-characterized in medical question answering. Most high-visibility medical LLM studies have focused either on very large models or on proprietary systems, whereas the trade-offs among predictive performance, inference efficiency, model size, and stability in smaller open models are less systematically documented [3, 8]. This gap is especially important in scenarios where local execution, hardware constraints, cost control, and reproducibility are first-order concerns. Our study is positioned in this space: rather than competing with frontier proprietary systems, we focus on benchmarking lightweight open-source models under a unified and resource-conscious setting.

### 2.4 Trustworthiness, Reliability, and Responsible Deployment

A growing body of work argues that evaluation in healthcare should be grounded in trustworthiness rather than raw benchmark performance alone. Surveys on medical LLMs and trustworthiness in healthcare repeatedly identify key dimensions including truthfulness, privacy, safety, fairness, robustness, explainability, and regulatory alignment [1, 8]. Likewise, commentary and review papers have stressed that benchmark scores must be interpreted alongside broader considerations of clinical responsibility, possible harm, and deployment governance [2, 3].

This perspective has two implications for the present study. First, lightweight model benchmarking should be interpreted as an investigation of capability boundaries rather than evidence of clinical readiness. Second, evaluation should be broadened to include efficiency, inter-model agreement, and subject-level heterogeneity, all of which contribute to a more realistic understanding of model behavior in healthcare contexts. In this sense, our work complements prior medical QA research by emphasizing reproducible benchmarking of lightweight open-source LLMs together with multi-view analysis of their performance and limitations.

## 3 Methods

### 3.1 Study Design

We designed MedScope as a lightweight and reproducible evaluation framework for benchmarking open-source large language models on medical multiple-choice question answering. The overall workflow consisted of four stages: dataset preparation, model inference, metric-based evaluation, and multi-view visualization analysis. In contrast to studies that focus primarily on a single aggregate benchmark score, our framework emphasizes a more comprehensive characterization of model behavior, including predictive performance, inference efficiency, response validity, subject-level variability, and inter-model consistency [3, 8, 9].

### 3.2 Dataset and Sampling Strategy

We used MedMCQA as the benchmark source for this study. MedMCQA is a large-scale medical multiple-choice question answering dataset constructed from Indian medical entrance examinations and contains more than 194,000 questions spanning 21 medical subjects and thousands of topics [9]. Since the purpose of our work was to evaluate lightweight open-source models under a resource-conscious but still informative setting, we did not use the full benchmark. Instead, we sampled 1,000 questions from the validation split with a fixed random seed to ensure reproducibility.

For each sampled instance, we retained the question stem, four candidate options, the gold answer, the subject label, and the topic label. Questions with incomplete option fields or invalid answer mappings were excluded during preprocessing. This strategy ensured that all evaluated samples had a consistent four-option structure suitable for unified prompting and automatic scoring.

### 3.3 Evaluated Models

We evaluated six lightweight open-source LLMs from three representative model families: LLaMA, Qwen, and Gemma. Specifically, the compared models were `llama3.2:1b, llama3.2:3b, qwen2.5:1.5b, qwen2.5:3b, gemma3:1b`, and `gemma3:4b`. These model families were selected because they are among the most visible open-weight LLM series and provide multiple lightweight parameter scales suitable for local deployment and reproducible experimentation [5, 13, 16].

All experiments were conducted under a consistent evaluation protocol. The models were queried locally through the Ollama runtime on macOS, which allowed us to compare them under the same hardware and software environment. We intentionally focused on lightweight variants rather than frontier large-scale models, because our goal was not to maximize absolute benchmark performance, but to analyze the trade-offs among performance, efficiency, and stability in practically deployable open-source models [3, 8].

### 3.4 Prompting Protocol

To minimize confounding introduced by prompt engineering, we adopted a unified prompt template for all models. Each question was formatted as a standard four-option multiple-choice item, and the model was instructed to return only one answer option from A, B, C, or D. The prompt was designed to constrain the output space and simplify automatic extraction of predictions.

Formally, given a question *q* and four candidate options {*o*_*A*_, *o*_*B*_, *o*_*C*_, *o*_*D*_}, the prompting objective was to obtain a single predicted label

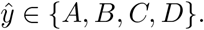

If the raw model output did not directly match one of these four labels, a rule-based post-processing procedure was applied to extract the final prediction from patterns such as `Answer: A`. Outputs that still could not be mapped into the valid answer space were marked as invalid.

### 3.5 Evaluation Metrics

We evaluated model behavior from four primary quantitative dimensions: accuracy, macro-F1, invalid response rate, and average inference time per question.

#### Accuracy

Accuracy measures the proportion of correctly answered questions:

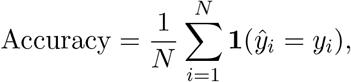

where *N* denotes the number of evaluated questions, *y*_*i*_ is the gold label, and ŷ_*i*_ is the predicted label for the *i*-th sample.

#### Macro-F1

Because the answer labels are categorical and class balance may vary in the sampled subset, we additionally report macro-F1. For each class *c* ∈ {*A, B, C, D*}, precision and recall are computed as

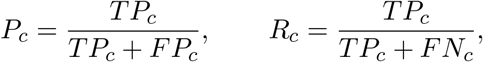

and the class-specific F1 score is

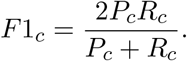

Macro-F1 is then obtained by averaging over all four classes:

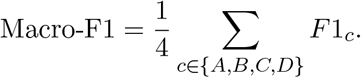

#### Invalid response rate

To quantify output controllability under the constrained prompting setup, we define invalid response rate as

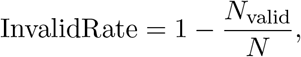

where *N*_valid_ is the number of predictions that can be successfully mapped to one of the four legal answer labels.

#### Inference efficiency

To reflect deployment-oriented efficiency, we record the average inference time per question:

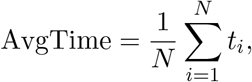

where *t*_*i*_ is the wall-clock response time for the *i*-th sample.

### 3.6 Subject-wise and Agreement Analysis

In addition to overall metrics, we analyzed performance differences across medical subjects by grouping samples according to the `subject_name` field provided by MedMCQA. Subject-wise accuracy was used to examine whether a model’s competence is evenly distributed or concentrated in particular domains. This analysis is important because aggregate performance alone may obscure substantial heterogeneity across specialties [8, 9].

We also measured pairwise prediction agreement between models. For two models *m*_*a*_ and *m*_*b*_, agreement was defined as

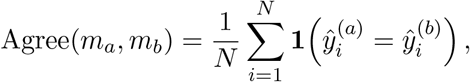

where 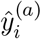 and 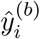 denote the predictions of the two models on sample *i*. This metric does not measure correctness directly; rather, it captures the extent to which models behave similarly in their decision patterns.

### 3.7 Visualization Strategy

To support more interpretable analysis, we supplemented metric tables with multiple visualization views. Clustered heatmaps were used to show subject-wise performance patterns and reveal cross-model similarity structures. Pareto-style trade-off plots were used to jointly visualize efficiency and predictive performance. Radar charts summarized multi-metric profiles for each model, while confusion matrices were used to inspect option-level prediction bias. Finally, agreement matrices and multi-panel dashboards were constructed to provide a compact but comprehensive summary of model behavior.

### 3.8 Implementation Details

All experiments were implemented in Python. Dataset preprocessing, inference logging, metric computation, and visualization were conducted through a unified local evaluation pipeline. To improve reproducibility, the sampling seed, prompt template, and post-processing logic were fixed across all models. The resulting framework is lightweight, transparent, and suitable for repeated benchmarking of open-source LLMs under controlled settings.

## 4 Experiments

### 4.1 Experimental Setup

We evaluated six lightweight open-source large language models from three representative model families: LLaMA, Qwen, and Gemma. The compared models were `llama3.2:1b, llama3.2:3b, qwen2.5:1.5b, qwen2.5:3b, gemma3:1b`, and `gemma3:4b`. These models were selected because they are openly available, lightweight enough for local deployment, and representative of current open-weight model families [5, 13, 16].

The evaluation was conducted on a sampled subset of 1,000 questions derived from MedM-CQA, where each instance contains a question stem, four answer options, a gold label, and associated subject metadata such as `subject_name` and `topic_name`.

Following prior benchmark-oriented work in medical LLM evaluation, we report both predictive and deployment-relevant metrics, including accuracy, macro-F1, invalid response rate, and average inference time per question [3,8,9]. Accuracy and macro-F1 capture answer quality, invalid response rate reflects output controllability under constrained prompting, and average inference time serves as a lightweight proxy for deployment efficiency.

### 4.2 Overall Results

Table 1 summarizes the evaluated models and their corresponding families, while Table 2 reports the main experimental results on the 1,000-question benchmark subset. Several observations can be made.

**Table 1.** Overview of the evaluated lightweight open-source LLMs.

| Model | Family | Scale | Deployment Setting |
| --- | --- | --- | --- |
| llama3.2:1b | LLaMA | 1B | Local lightweight inference |
| llama3.2:3b | LLaMA | 3B | Local lightweight inference |
| qwen2.5:1.5b | Qwen | 1.5B | Local lightweight inference |
| qwen2.5:3b | Qwen | 3B | Local lightweight inference |
| gemma3:1b | Gemma | 1B | Local lightweight inference |
| gemma3:4b | Gemma | 4B | Local lightweight inference |

**Table 2.** Main results on the 1,000-question MedMCQA subset. Best values are in **bold**; second-best values are underlined. Lower is better for Invalid Rate and Avg. Time.

| Model | Accuracy | Macro-F1 | Invalid Rate | Avg. Time (s) |
| --- | --- | --- | --- | --- |
| llama3.2:3b | **0.2755** | <u>0.1851</u> | 0.1470 | 0.8695 |
| gemma3:4b | <u>0.2530</u> | **0.2076** | **0.0000** | 0.7092 |
| qwen2.5:1.5b | 0.2150 | 0.1840 | **0.0000** | **0.1712** |
| llama3.2:1b | 0.2144 | 0.1785 | 0.0110 | 0.7010 |
| gemma3:1b | 0.2020 | 0.1729 | **0.0000** | <u>0.2632</u> |
| qwen2.5:3b | 0.2000 | 0.1716 | **0.0000** | 0.2691 |

**Table 3.** Family-level average performance across evaluated model scales.

| Family | Accuracy | Macro-F1 | Invalid Rate | Avg. Time (s) |
| --- | --- | --- | --- | --- |
| LLaMA | **0.2450** | 0.1818 | 0.0790 | 0.7852 |
| Gemma | <u>0.2275</u> | **0.1902** | **0.0000** | 0.4862 |
| Qwen | 0.2075 | 0.1778 | **0.0000** | **0.2202** |

First, `llama3.2:3b` achieved the highest overall accuracy, reaching **0.2755**, which suggests that increasing parameter scale within the LLaMA family improved answer correctness under the present evaluation setting. However, this gain came at the cost of the highest invalid response rate (**0.1470**) and the slowest inference speed (0.8695 seconds per question), indicating a clear trade-off between predictive performance and output controllability.

Second, `gemma3:4b` demonstrated the strongest macro-F1 (**0.2076**) while maintaining a zero invalid response rate. Compared with `llama3.2:3b`, its accuracy was slightly lower (0.2530 vs. 0.2755), but its responses were more stable and better balanced across answer classes. This suggests that model quality in medical multiple-choice question answering should not be judged by accuracy alone.

Third, the Qwen family exhibited the strongest efficiency characteristics. In particular, `qwen2.5:1.5b` achieved the fastest inference speed (**0.1712** seconds per question) while also producing no invalid outputs. Although its accuracy was lower than the best-performing LLaMA and Gemma variants, it offered a strong efficiency–performance trade-off under resource-constrained evaluation.

Finally, the results reveal substantial heterogeneity across model families and scales. Larger models did not uniformly dominate smaller ones across all metrics, and the best-performing model varied depending on whether one prioritizes raw accuracy, class-balanced performance, output validity, or efficiency. These findings support the need for multi-dimensional evaluation in lightweight medical LLM benchmarking rather than relying on a single aggregate score [3, 8].

### 4.3 Family-Level Trends

To better understand broader architectural tendencies, we further examined family-level trends across LLaMA, Qwen, and Gemma. Averaged over the two evaluated scales, the LLaMA family achieved the highest mean accuracy, while Gemma showed the strongest balance between accuracy and macro-F1 together with consistently valid outputs. By contrast, Qwen yielded the fastest average inference speed among the three families, suggesting that different open-source model families occupy different regions of the performance–efficiency trade-off space. This family-level heterogeneity further justifies the use of multi-view evaluation and visual analysis in lightweight medical benchmarking.

## 5 Results Visualization and Qualitative Analysis

To complement the quantitative results reported in Table 2, we further conducted a multi-view visual analysis of model behavior. Rather than treating medical question answering performance as a single-number ranking problem, we examined how lightweight open-source LLMs differed across medical subjects, evaluation dimensions, inter-model agreement patterns, and prediction flow structures. These visual analyses provide a more interpretable understanding of model behavior and reveal patterns that are not immediately visible from aggregate scores alone.

### 5.1 Multi-metric Overview

Figure 1 presents a comprehensive multi-panel summary of the six evaluated models. Several general patterns can be observed. First, the models do not exhibit a uniform ordering across all dimensions. Although `llama3.2:3b` achieved the highest overall accuracy, it was not dominant in every aspect. In particular, `gemma3:4b` achieved the best macro-F1 and maintained fully valid outputs, while `qwen2.5:1.5b` exhibited the strongest inference efficiency. This confirms that lightweight medical LLM evaluation is fundamentally multi-objective: a model that performs best under one criterion may no longer be optimal when robustness, output controllability, or latency is taken into account.

**Figure 1.**
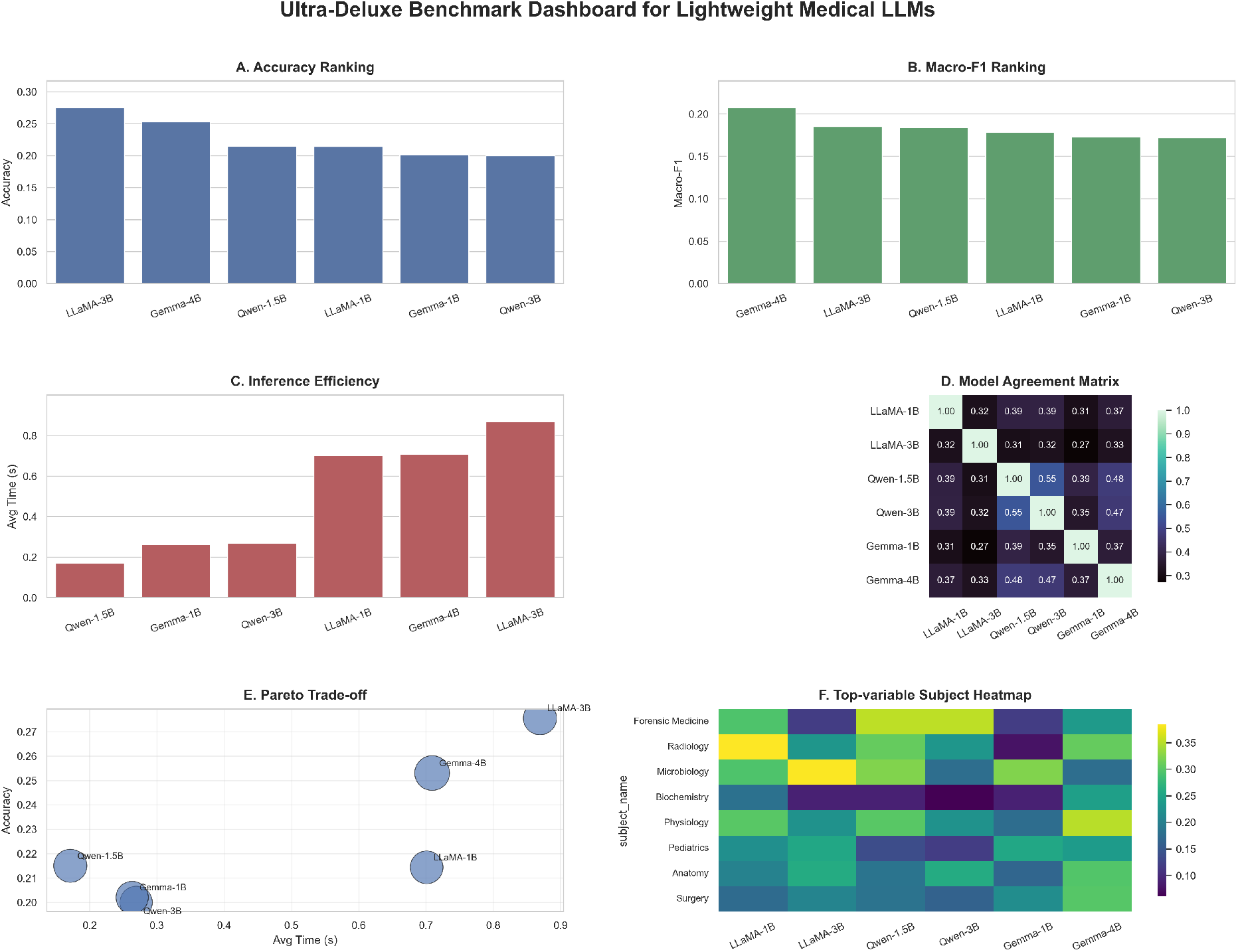
Comprehensive multi-panel summary of the six lightweight open-source LLMs, showing overall ranking patterns in accuracy, macro-F1, inference efficiency, agreement structure, and trade-off landscape.

Second, the visual ranking patterns suggest that model family effects remain substantial even at relatively small scales. The Gemma and LLaMA families tend to occupy stronger positions in overall predictive quality, whereas the Qwen family shows a clearer efficiency advantage. Such differences imply that lightweight medical QA performance is shaped not only by model size, but also by architectural family and training design.

### 5.2 Subject-wise Heterogeneity

Figure 2 shows the clustered heatmap of subject-wise accuracy across the six evaluated models. This figure reveals substantial heterogeneity across medical disciplines. Performance differences are not evenly distributed: some subjects, such as microbiology, forensic medicine, physiology, and radiology, display relatively large variation across models, whereas others exhibit more compressed differences. This indicates that model competence in medical question answering is not uniform across specialties, even when overall benchmark scores appear relatively close.

**Figure 2.**
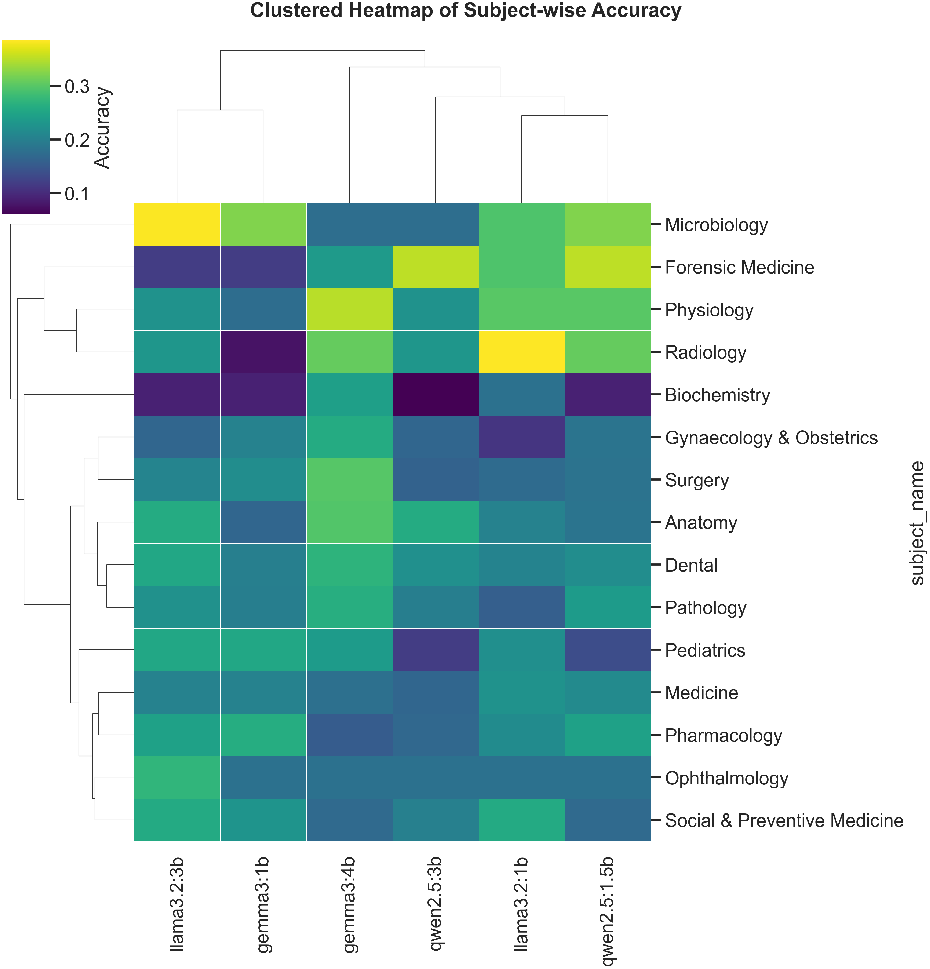
Clustered heatmap of subject-wise accuracy across the six lightweight models. The visualization reveals substantial heterogeneity across medical subjects and highlights similarity structures among models.

The clustered structure also provides useful insights into model similarity. Models from related performance regimes tend to group together, suggesting that subject-wise behavioral patterns may be more informative than aggregate accuracy alone. In particular, the higher-capacity variants tend to cluster closer to each other in several subject domains, while lighter models show more uneven or fragmented subject-level profiles. These results reinforce the importance of subject-aware benchmarking in medical AI, since a single global score can conceal meaningful weaknesses in particular domains.

### 5.3 Efficiency–Performance Trade-off

The Pareto-style bubble plot in Figure 3 visualizes the trade-off between average inference time and predictive accuracy, with bubble size reflecting an additional quality-related metric. This figure makes the deployment-oriented tension particularly clear. The Qwen models, especially `qwen2.5:1.5b`, occupy the low-latency region, demonstrating their practical advantage in fast-response scenarios. In contrast, `llama3.2:3b` reaches the strongest accuracy but lies in the slowest region of the trade-off space. The Gemma models occupy an intermediate zone, with `gemma3:4b` representing a relatively balanced point between effectiveness and efficiency.

**Figure 3.**
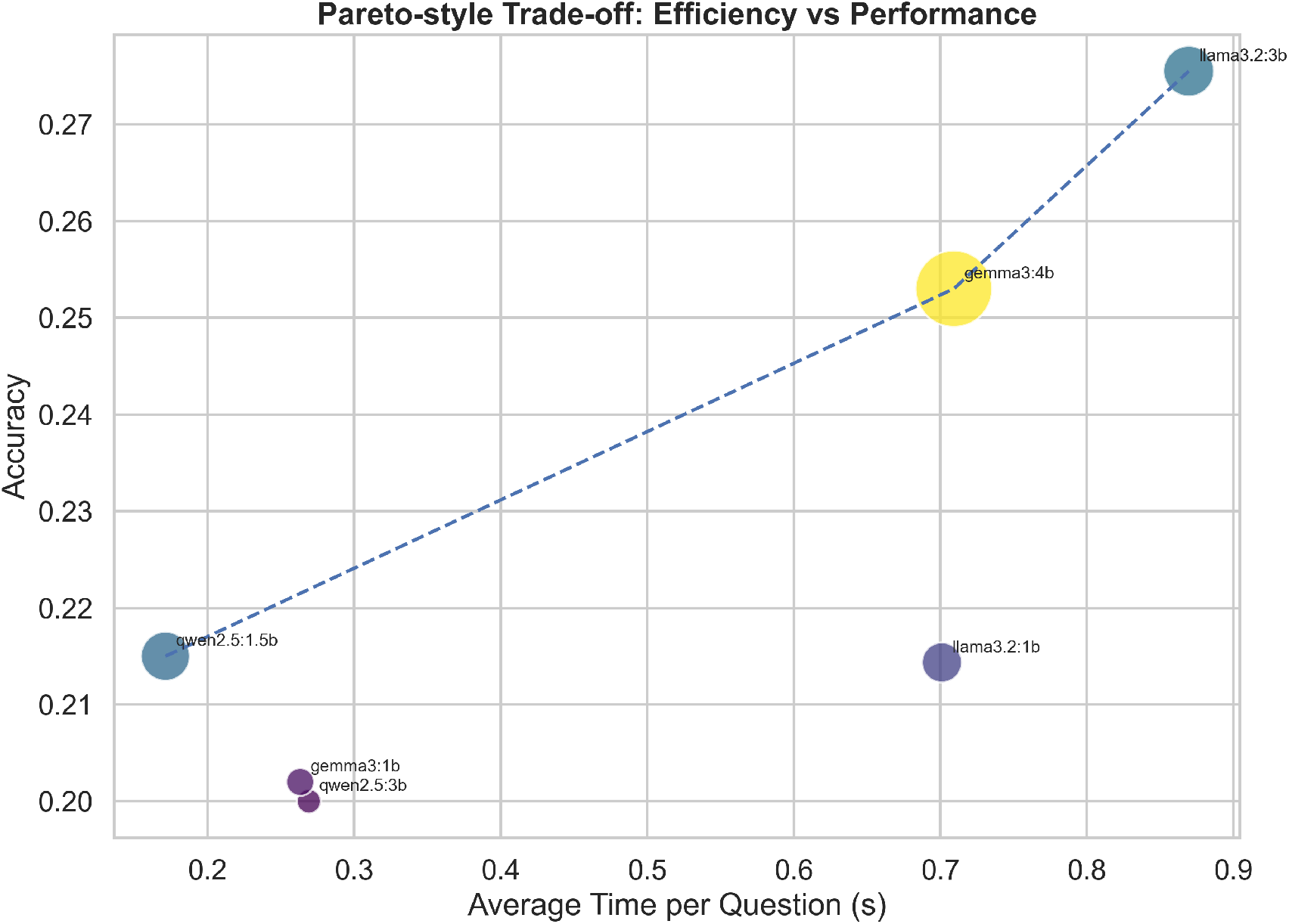
Pareto-style trade-off between predictive performance and inference efficiency. Each point denotes a model, and the relative bubble size reflects an additional performance-related dimension.

This trade-off perspective is important for realistic deployment considerations. In many resource-constrained settings, the most accurate model may not be the most useful if inference cost, latency, or operational stability are critical. Conversely, the fastest model may sacrifice too much predictive quality. The Pareto-style view therefore provides a more informative decision surface than a single leaderboard ranking.

### 5.4 Agreement and Behavioral Similarity

Figure 4 presents the pairwise prediction agreement matrix across models. A notable pattern is that models from the same family or similar scale tend to exhibit relatively stronger agreement, whereas cross-family agreement is generally lower. In particular, the two Qwen variants show comparatively high mutual agreement, suggesting that their decision patterns are more internally consistent than their agreement with other families.

**Figure 4.**
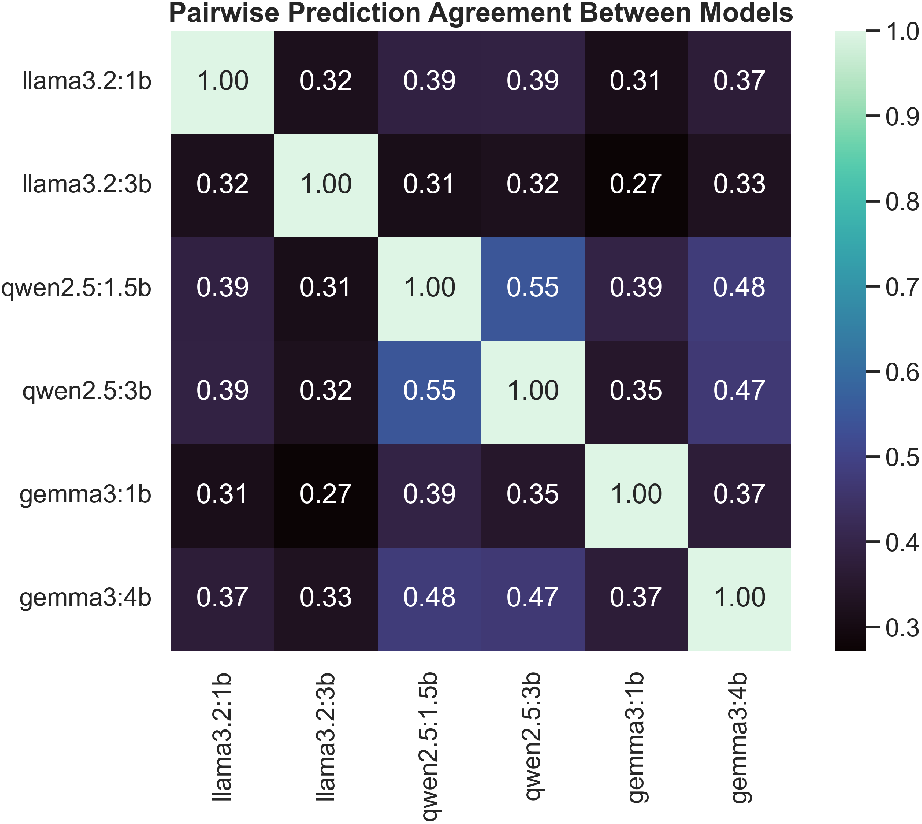
Pairwise prediction agreement between models. Higher agreement indicates stronger similarity in decision patterns, but does not necessarily imply higher correctness.

Importantly, agreement should not be interpreted as correctness. Instead, it reflects behavioral similarity: two models may agree because they are both correct, or because they make similar errors. Nevertheless, this matrix is still highly informative, as it reveals the extent to which different open-source LLMs rely on similar decision tendencies. The relatively moderate agreement values across many model pairs suggest that lightweight medical LLMs preserve meaningful diversity in their outputs, which may be useful for future ensemble-style or committee-based evaluation.

To further emphasize this structure, Figure **??** visualizes the agreement relationships as a network. Models connected by stronger edges tend to form local similarity clusters, offering a visually intuitive summary of inter-model behavior. This representation is especially useful when discussing whether different open-source model families provide complementary rather than redundant predictions.

### 5.5 Multi-dimensional Profile Comparison

Figure **??** provides a radar-style comparison over multiple normalized dimensions, including accuracy, macro-F1, speed, and reliability. This figure is useful for identifying each model’s characteristic profile rather than simply ranking models from best to worst. For example, `gemma3:4b` exhibits a broader and more balanced profile, while `qwen2.5:1.5b` is more skewed toward efficiency. In contrast, `llama3.2:3b` shows its strongest advantage in predictive performance but a less favorable efficiency contour.

Such multi-dimensional comparison is particularly valuable in medical settings, where different deployment scenarios may prioritize different objectives. Educational support systems, edge deployment, and local experimentation may value efficiency and validity more strongly, whereas research benchmarking may prioritize absolute predictive quality. The radar view therefore helps contextualize model suitability across different use cases.

### 5.6 Prediction Flow and Error Structure

To gain a more qualitative understanding of model error patterns, we further visualize the flow from gold labels to predicted labels using a Sankey-style diagram (Figure 5). This figure highlights how correct and incorrect predictions are distributed across answer classes. Rather than reducing all errors to a single scalar, the flow structure shows whether a model exhibits asymmetric confusion tendencies, such as a stronger attraction toward a specific option label. This is particularly useful in multiple-choice evaluation, where option-level bias can remain hidden under aggregate accuracy and macro-F1.

**Figure 5.**
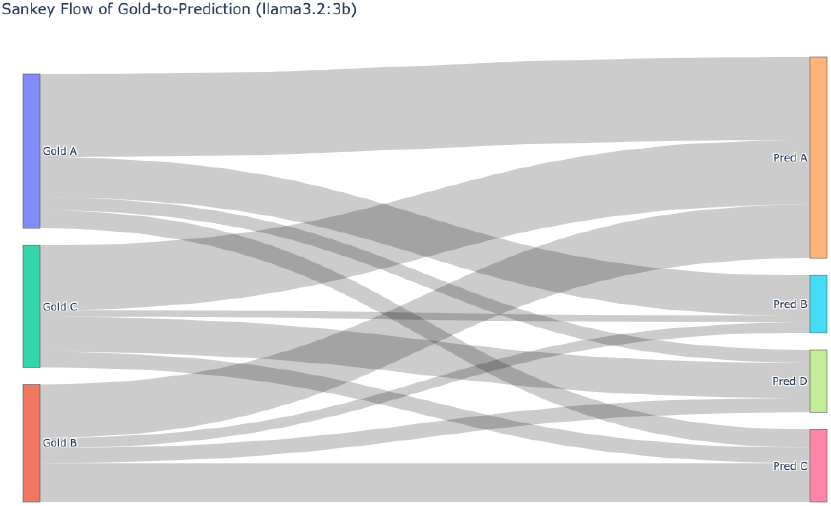
Sankey-style visualization of the gold-to-prediction flow for the strongest-performing model, illustrating the distribution of correct predictions and option-level confusion patterns.

Together with the confusion matrices shown in the supplementary visual set, the Sankey-style visualization suggests that some models make more concentrated misclassification patterns, whereas others distribute their errors more diffusely across answer options. This distinction is important because structured error patterns may be easier to diagnose and potentially mitigate through prompt refinement or decoding constraints.

### 5.7 Summary of Visual Findings

Overall, the visual analyses consistently support three conclusions. First, lightweight open-source medical LLMs exhibit substantial heterogeneity across model families, scales, and subject domains. Second, the trade-off between predictive quality and inference efficiency is non-trivial and must be explicitly considered in deployment-oriented evaluation. Third, aggregate accuracy alone is insufficient to characterize model behavior in medical QA, as subject-level variability, agreement structure, and error flow provide additional evidence about robustness and practical limitations.

These findings strengthen the central claim of this work: benchmarking lightweight medical LLMs should be approached as a multi-dimensional evaluation problem rather than a single-score comparison task.

## 6 Discussion

This study provides a lightweight but multi-dimensional benchmark of open-source LLMs for medical multiple-choice question answering. Several important findings emerge from our experiments. First, lightweight models from different families occupy distinct regions of the performance–efficiency space. While `llama3.2:3b` achieved the highest overall accuracy, `gemma3:4b` showed the strongest macro-F1 together with fully valid outputs, and `qwen2.5:1.5b` demonstrated the strongest inference efficiency. These results suggest that lightweight medical LLM evaluation should not be reduced to a single score. Instead, different models may be preferable under different deployment priorities, including predictive quality, response validity, latency, or class-balanced behavior [3, 8].

Second, our subject-wise analyses reveal that the capability of lightweight medical LLMs is highly uneven across disciplines. Aggregate benchmark scores alone obscure important variation between subject domains, as some specialties exhibit substantially larger cross-model gaps than others. This observation is consistent with broader concerns in the medical LLM literature that benchmark-level success does not imply uniform competence across real healthcare tasks [8, 11]. In practical terms, this means that even relatively competitive lightweight models should not be interpreted as broadly reliable medical assistants without additional task- and domain-specific validation.

Third, the agreement analysis highlights that model similarity and model correctness are not equivalent. Higher pairwise agreement may reflect shared strengths, but it may also reflect shared error tendencies. Conversely, moderate agreement across model families suggests that open-source lightweight LLMs retain meaningful behavioral diversity. This may prove useful for future research on model ensembles, committee-style verification, or disagreement-aware medical AI systems, where diversity itself can become a source of robustness.

From a broader healthcare perspective, our findings support a cautious interpretation of lightweight open-source LLMs. Their accessibility, transparency, and local deployability make them highly attractive for reproducible experimentation and low-resource research settings [5, 13, 16]. However, the current results remain far from supporting autonomous clinical deployment. In high-stakes medical contexts, even moderate prediction error can be consequential, and performance variability across specialties further increases uncertainty. This reinforces the view that medical LLMs should presently be treated as assistive tools rather than independent decision-makers [1, 15].

### 6.1 Implications for Responsible Deployment

The practical relevance of lightweight open-source models lies not only in their benchmark performance, but also in their potential role in privacy-conscious and resource-constrained environments. Compared with proprietary cloud-based systems, open-weight lightweight models are easier to study, reproduce, and execute locally, which may reduce data-sharing barriers and improve transparency in experimental design [8]. At the same time, local deployability should not be conflated with safety. Responsible use in healthcare requires explicit awareness of model limitations, human oversight, clear accountability boundaries, and context-specific validation before any downstream use in patient-facing or clinically consequential settings [6, 15].

### 6.2 Limitations

This study has several limitations. First, the evaluation was conducted on a sampled subset of MedMCQA rather than the full benchmark. Although this design is appropriate for lightweight and reproducible experimentation, it may limit the statistical stability and external validity of some findings. Second, our benchmark focuses on multiple-choice medical question answering, which is only one aspect of medical intelligence. It does not capture broader capabilities such as clinical dialogue, longitudinal reasoning, uncertainty communication, or interaction with multimodal patient data [11, 14].

Third, our prompting strategy intentionally emphasized simplicity and comparability rather than prompt optimization. As a result, the reported results should be interpreted as behavior under a controlled lightweight benchmark rather than the maximal achievable performance of each model. Fourth, we evaluated only lightweight open-source models and did not compare them with frontier proprietary systems. This was a deliberate design choice motivated by reproducibility and deployment realism, but it also means that our findings characterize relative differences within the lightweight open-source regime rather than the broader frontier of medical LLM capability.

Finally, our study remains benchmark-oriented. Even though we extended the analysis beyond aggregate accuracy through subject-wise, agreement-based, and visualization-driven perspectives, benchmark performance is still not equivalent to clinical utility. Real-world deployment would require prospective validation, clinician-in-the-loop workflows, safety evaluation, and governance mechanisms that go beyond the scope of the present work [1, 2, 15].

## 7 Conclusion and Future Work

In this work, we presented MedScope, a lightweight benchmark for evaluating open-source large language models on medical multiple-choice question answering. Using 1,000 sampled questions from MedMCQA, we systematically compared six lightweight models from the LLaMA, Qwen, and Gemma families under a unified evaluation protocol. Beyond conventional benchmark metrics, we incorporated inference efficiency, invalid response rate, subject-wise variability, inter-model agreement, and multiple visualization-based analyses. Together, these results show that lightweight open-source medical LLMs exhibit meaningful but highly uneven capabilities, with clear trade-offs among predictive performance, efficiency, and behavioral stability.

Our study makes two broader contributions. First, it demonstrates that lightweight open-source medical LLMs can be evaluated in a reproducible and practically deployable setting without relying on closed commercial systems. Second, it shows that medical LLM benchmarking should move beyond single-metric comparison and toward a more multi-dimensional framework that better captures real model behavior. In this sense, MedScope provides not only a benchmark result set, but also an analysis perspective for understanding lightweight models in healthcare-related tasks.

There are several promising directions for future work. One important extension is to evaluate lightweight open-source models on broader medical reasoning settings beyond multiple-choice QA, including open-ended question answering, conversational diagnosis, uncertainty-aware response generation, and longitudinal clinical reasoning [11, 14]. Another direction is to investigate whether ensemble strategies, verifier models, or disagreement-aware pipelines can exploit behavioral diversity across model families to improve robustness. It would also be valuable to study how prompt design, decoding strategies, or retrieval augmentation affect the performance–efficiency trade-off in lightweight medical LLMs.

More fundamentally, future work should connect benchmark-centered evaluation with real deployment constraints. This includes clinician-in-the-loop assessment, prospective workflow studies, privacy-preserving local deployment, and stronger safety-governance frameworks for medical AI [1,6,15]. We hope that MedScope can serve as a useful baseline for these future efforts and contribute to more transparent, reproducible, and responsible development of lightweight LLMs in medicine.

## Data Availability

All data produced in the present study are available upon reasonable request to the authors

